# Glaucoma classification through SSVEP derived ON- and OFF-pathway features

**DOI:** 10.1101/2024.08.22.24312443

**Authors:** Martin T. W. Scott, Hui Xu, Alexandra Yakovleva, Robert Tibshirani, Jeffrey L. Goldberg, Anthony M. Norcia

**Affiliations:** Department of Psychology, Stanford University; Department of Statistics, Stanford University; Spencer Center for Vision Research, Byers Eye Institute, Department of Ophthalmology, Stanford University; Wu-Tsai Neurosciences Institute, Department of Psychology, Stanford University

## Abstract

Recent evidence from small animal models and human electrophysiology suggests that the OFF-pathway is more vulnerable to glaucomatous insult than the ON-pathway. Thus, OFF-pathway based measurements of visual function may be useful in the diagnosis of Glaucoma. The steady-state visually evoked potential (SSVEP) can be used to non-invasively make such functional measurements. Here, we examine whether OFF- and ON-pathway biasing SSVEP measurements differently predict glaucoma diagnosis using a large cohort of 98 glaucoma patients and 71 controls. Using both a logistic regression with k-fold cross-validation and a random forest classifier, we show that OFF-pathway biasing features produce a small improvement in predictive accuracy over ON-pathway biasing features. However, despite our inclusion of many more response features and the retention of both participants’ eyes, our classifier did not perform as well as previous reports that used the isolated-check VEP. This is likely a result of the relatively small amount of data we collected for each participant, but may also be explained by the absence of any train-test splitting in preexisting work. Nevertheless, our results support further exploration of the diagnostic potential of OFF-pathway biasing functional biomarkers for glaucoma.

## Introduction

Recent work in mouse models of glaucoma has suggested that OFF retinal ganglion cells (RGCs) are more susceptible to damage than ON RGCs (Della Santina et al., 2013; El-Danaf & Huberman, 2015; Ou et al., 2016; Puyang et al., 2017). In human glaucoma patients, these reports have been corroborated by non-invasive electrophysiolical measures of ON and OFF pathway function (Horn et al., 2011; Kong et al., 2021; Norcia et al., 2022; Pangeni et al., 2012). These findings are also reflected in retinal imaging of the ON- and OFF-sub-laminae of the Inner Plexiform Layer (IPL) (Ghassabi et al., 2022). In each of these cases, evidence for preferential OFF-pathway damage was found using group-level statistical analyses, which is valuable for the directing further exploration of biomarkers for glaucoma. However, an important future application of sub-laminae specific imaging and electrophysiology is the detection of glaucomatous field loss or progression in individual patients.

Several electrophysiological studies have addressed individual patient classification using the isolated-check visually-evoked potential (icVEP) a form of Steady-State VEP (X.-W. Chen & Zhao, 2017; X. Chen & Zhao, 2017; Fan et al., 2018; Kolomeyer et al., 2020; L. J. Xu et al., 2017; L.-j. Xu et al., 2020; Zemon et al., 2008). The icVEP protocol has variations that are designed to bias neural responses towards either the ON or OFF pathway. This biasing is accomplished by presenting a rectilinear grid of small patches that sinusoidally modulate their luminance either above the luminance of an otherwise uniform background (ON-pathway favoring) or below it (OFF-pathway favoring). More recent work using this paradigm in glaucoma has focused on ON-favoring stimuli, based on early group-level and individual-level analyses showing greater effects of glaucoma with incremental sinusoidal stimulation (Greenstein et al., 1998; Zemon et al., 2008). More recent SSVEP work used saw-tooth modulation (Norcia et al., 2022), which has the potential to more effectively bias responses towards either pathway (Kremers et al., 1993). Given recent SSVEP and ERG group-level results suggesting greater OFF-pathway loss in glaucoma, here we re-visit the relative accuracy of binary classification (patient vs control) of individuals on the basis of on ON- and OFF-pathway biasing SSVEPs in a large sample of patients and controls.

Our approach differs from previous classification-based SSVEP studies in two principle ways in addition to the use of sawtooth vs sinsusoidal stimulation. We recorded 128-channel EEG rather than single channel EEG and used spatial filtering for dimension reduction and signal enhancement. Spatial filtering was accomplished via an objective dimension reduction approach that weights channels based on a fundamental aspect of the SSVEP: channels with stimulus-locked responses are the channels of interest (Dmochowski et al., 2015). Secondly, when training two-group classifiers, we used a cross-validation method that avoids over-fitting and simultaneously allows for the inclusion of both participant’s eyes (where available) controlling for the dependency structure imposed by having multiple eyes from an individual in the dataset. Our results highlight the importance of considering dependency structure when assessing classification performance and, importantly, indicate that classification accuracy is higher and more strongly driven by OFF-derived response features over ON-derived response features.

## Methods

### Participants

Experiments proceeded after approval by the Institutional Review Board of Stanford University. Written informed consent was obtained from all participants, and all research conformed to the tenets of the Helsinki accord for the use of human participants. The participants were recruited from the Stanford University community or were patients at glaucoma/optometry clinics of the the Byers Eye Institute at Stanford University. Participants included a group of 98 adults (mean age = 59.3, 39 females, 54 males, 5 unknown) with glaucoma and a group of 71 adults with similar age and gender (mean age = 61.7, 35 females, 29 males, 7 unknown) without glaucoma or other ocular pathology. Data from 177 eyes of 98 patients and 140 eyes of 71 controls was used in the classification analysis.

Glaucoma patient inclusion criteria included the diagnosis of glaucoma, best corrected visual acuity of 20/70 or better in the study eye(s), absence of other ophthalmic issues that may impact vision, and cognitive abilities sufficient to participate in the study. Glaucoma was diagnosed basis of glaucomatous optic nerve head damage and retinal nerver fiber layer thinning via fundoscopy and optical coherence tomography (respectively) and typical visual field loss on the 24-2 Humphrey visual field. Typical field loss was defined as a positive glaucoma hemifield test or a cluster of at least three points below p = 0.05, with at least one point below p = 0.01. Humphrey visual field testing was performed for all patients with glaucoma on the day of or within 3 months prior to the VEP recording. Group-level data from 61 of our patients and 37 control participants have been reported previously (Norcia et al., 2022). The distributions of mean deviations (MDs) and ages of the patient sample are shown in Figure 1. For MD correlation (Figure 1, right panel) 72 patients with both OD and OS measurements are included, and a regression line was using the “fitlm()” function in MATLAB 2022b.

**Figure 1.**
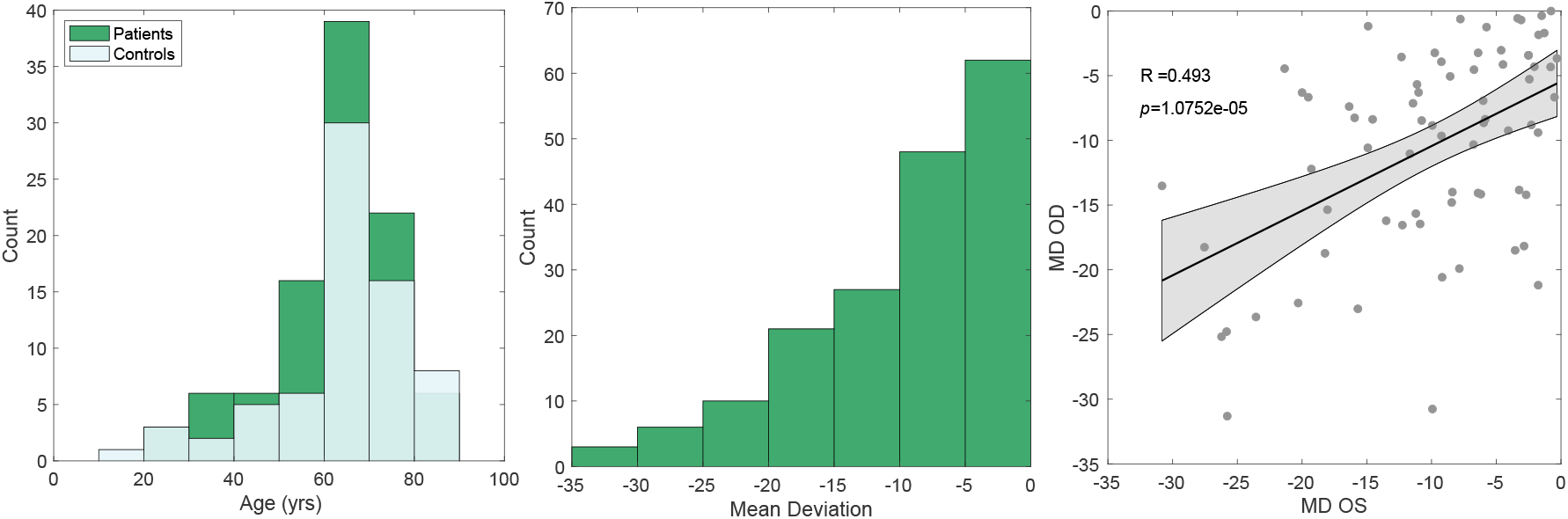
Demographic information of participants. The left panel contains the distribution of ages for patients and controls. The middle panel contains the distribution of mean deviation (MD) values for patients. The right panel contains the left-eye (OS) MD scores plotted against the right-eye (OD) MD scores, and a the fit of a linear regression (shaded region = 95% CI on the fit).

### Visual Stimuli

Responses to ON- and OFF-pathway biasing stimuli were measured using an hexagonal array of flickering probes. The entire array subtended 42°× 25° of visual angle. Two types of hexagonal elements are present in the stimulus array: probes and pedestals. Pedestal hexagons are larger elements that have a fixed luminance of 47 cd/m2, while probe hexagons are smaller elements within pedestals with luminance modulation that was experimentally manipulated. All hexagons were presented against a low-luminance background of 11 cd/m2 (see panel B of Figure 2). Elements that straddled the horizontal and vertical meridian were eliminated (see Figure 2C for a schematic example). Probe elements were temporally modulated according to a sawtooth profile, the fast-phase of which was set to bias evoked responses either towards the ON or OFF pathway (Kremers et al., 1993). ON-pathway biasing stimuli were defined as probes that rapidly increased in luminance and slowly decreased, while OFF pathway biasing stimuli were defined as probes that rapidly decreased in luminance and slowly increased (see wave-forms in Figure 2A). All stimulus elements were scaled with eccentricity as detailed previously (Norcia et al., 2020). Stimuli were presented on a SONY PVM-2541 monitor (1920 x 1080 pixels) and viewed monocularly at a distance of 70 cm. Display pipeline delays and EEG pipeline delays were measured with a photocell and have been corrected. All probes in the array flickered at 2.727 Hz synchronously with an identical temporal waveform. SSVEPs were measured using probes of 20% contrast, with contrast defined using the Weber definition: 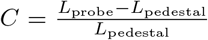, where *L* is the luminance of the probe or the pedestal.

**Figure 2.**
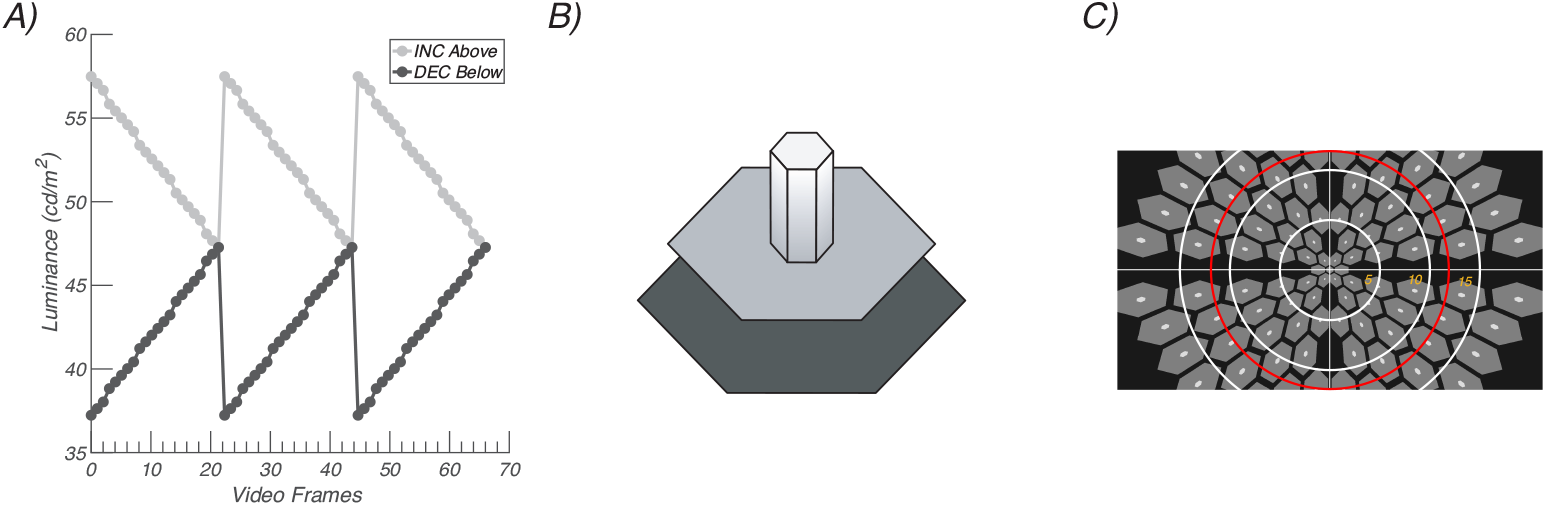
Visual stimulation paradigm. A) Sawtooth stimulation profiles for probes, with incremental probes presented above the pedestal luminance and decremental probes below. B) Probe-on-pedestal display element. The sawtooth-modulated probes (small white element rising from the gray pedestal) was sawtooth modulated, here above the pedestal. The probe was 20% of the size of the pedestal. The pedestal was surrounded by a larger black background. C) Scaled stimulus array. White rings indicate 5 deg increments of eccentricity from central fixation. The red ring is 12 deg in radius.

### Procedure

Participants viewed ON- and OFF-pathway biasing stimuli monocularly with their contra-lateral eye covered with an opaque patch. The trials were blocked by eye. Within an eye-testing block, increment and decrement trials were presented in random order. Trials lasted up to 13.2 sec with 3000 + 500 ms inter-trial intervals. Participants were instructed to withhold blinking and to fixate on the center element. For the conditions of present interest, participants saw 10 trials per condition, per eye (i.e. 40 trials total 2 pathways x 2 eyes x 10 trials). To control participant vigilance and clamp attention at a more constant value, a simple fixation task was presented concurrently during steady-state stimulation. Note, the conditions being analysed here are a subset of 12 total conditions the participants saw during a session. The other conditions were designed to optimise/examine unrelated stimulus parameters.

### EEG recording and artifact rejection

The EEG was recorded over 128 channels using Hydrocell SensorNets and NetStation 5.2 software (Electrical Geodesics, Eugene, OR). Prior to recording, individual electrodes were adjusted so that the impedance values were lower than 100 kΩ. The raw EEG was amplified (gain = 1000 at 24-bit resolution) and digitally filtered with a 0.3 Hz to 50 Hz band-pass filter. The data was then processed using in-lab software written in Objective C as follows. First, consistently noisy individual channels were detected, rejected, and substituted with the average of the six nearest-neighbour channels. Channels were classified as consistently noisy if over 15% of samples exceeded 30 µV (excluding breaks). After this, the data were re-referenced to the common average. Second, the 13.2-second trials were binned into 12 bins of 1.1 seconds. The first and last bin of each trial were always discarded. Third, to reject data containing coordinated muscle movements and blinks, 1.1s bins were excluded for all channels if more than 5% of channels exceeded an amplitude threshold of 60 µV. Fourth, 1.1s bins of individual channels were excluded if more than 10% of samples exceeded 30 µV. These light-touch artefact rejection criteria were derived empirically for adults over hundreds of previous recordings. They readily pick out muscle and blink artifacts as well as electrode motion artifacts which are not of neural origin, leaving relatively clean EEG. As one of our objectives was to realistically examine the clinical diagnostic potential of the SSVEP, we opted not to reject patients’ or controls’ data wholesale on the basis of signal strength or data quality metrics.

### Spectral Analysis

Spectral analysis was performed for each participant, for every sensor, at every 1.1s bin using a recursive least squares (RLS) filter (Tang & Norcia, 1995). Briefly, RLS is equivalent to the discrete Fourier transform (DFT) but is more effective when short trial lengths are used. Conceptually, RLS directly fits sine and cosine waves at selected stimulus-relevant frequencies to a time-series. This process yields complex-valued estimates of harmonic amplitudes that can be used in the same way as the amplitudes from the DFT. In the present work, RLS was performed up to the 4th harmonic of the stimulus frequency. Assuming a participant had no bins rejected, this yielded 80 spectral estimates per condition, per eye (10 bins x 10 trials x 4 harmonics). Note, that we use an “xF” nomenclature when referring to stimulus-related frequencies, such that “2F” refers to the 2nd harmonic of the stimulus frequency.

### Spatial filtering

Group-level reliable components analysis (RCA) in the frequency domain was used to reduce the dimensionality of the 128-channel data to a smaller number of more easily interpreted components, as previously detailed (Dmochowski et al., 2015). Briefly, each reliable component (RC) is a weighted sum of electrical potentials across all channels. The weight vectors are derived through an eigenvalue decomposition performed on a 128 x 128 matrix where each element represents the ratio of within-trial covariance (*R*_*xx*_) to cross-trial covariance (*R*_*xy*_). Solving this decomposition provides multiple ranked components (i.e. spatial filters) that maximise *R*_*xx*_*/R*_*xy*_, with the 1st RC containing the maximal contribution from channels with consistent cross-trial activity. This reflects a fundamental quality of the SSVEP, where repeated presentations of the same stimulus produce similar stimulus-locked neural activation across multiple trials. For classification, RCA spatial filters were trained at the group level for the control participants. Components were learned from the complex values of the first 4 harmonics of the stimulus frequency as higher harmonics had low signal-to-noise (SNR), even in control eyes (Norcia et al., 2020; Norcia et al., 2022). The raw data for both patients and controls were then projected through the control-derived weightings. We projected all participants’ data through the same weights to eliminate the possibility of differentiating the groups on the basis of a component sign-flip (due to the nature of eigenvalue decomposition). We consider the first 6 RCs for binary classification. The individual subject level data comprised their cross-trial vector mean for each harmonic and condition. Finally, individual complex-valued vector means were transformed into amplitude (*ρ*) and phase delay (*θ*) values. Phase delay is a circular variable, meaning it cannot be used directly for binary classification. So, we transformed it into the cos(*θ*) and sin(*θ*). This process yields the useful property of being able to separate features into amplitude-derived and phase-derived. We retained 6 RCA components and thus had 144 response features per eye (i.e. [cos(*θ*), sin(*θ*), *ρ*] over 4 harmonics x 6 RC components x two pathways).

### Case-control classification

To evaluate the diagnostic value of the ON and OFF-pathway SSVEP for glaucoma we trained a logistic regression classifier with elastic-net regularisation on our dataset of 169 individuals using the glmnet package (V4.18) (Friedman et al., 2021). We additionally trained a random forest classifier to produce estimates of feature importance using the randomForest package (V4.7-1.1) (Liaw & Wiener, 2007). We used 10-fold cross-validation (CV) to optimise the regularisation parameter of the elastic net and to produce robust performance metrics for both classification algorithms. Each individual was classified either as a patient or control. We trained classifiers using 144 VEP response features and two demographic features (sex and age). We employed two CV strategies for both the logistic regression and the random forest. In the first strategy, observations belonging to a given participant were always grouped together in the same CV fold, preserving the dependency structure, and preventing overly-optimistic performance estimates. We refer to this as the “paired eyes” strategy. For the second strategy, observations were randomly assigned to CV folds, such that the aforementioned dependency structure is not preserved. We refer to this as the “unpaired eyes” strategy. We evaluated classification performance on the basis of accuracy, sensitivity, specificity, and AUC (area under the receiver operating characteristic curve). For each error metric, we report the CV error, which is the recognized best-practice for estimating the prediction error of a model (Allen, 1974; Geisser, 1975; Stone, 1974).

## Results

We begin with a brief overview of the data used for classification. Figure 3 shows the scalp topography of the first 3 RCs for controls. The focal distribution of RC1 over posterior channels is consistent with responses originating from early cortical processing nodes, like primary visual cortex (V1), V2, and V3 (see the forward modeling work of Ales et al. (2010)). Reliable components 2 and 3 have more lateralized topographies, likely reflecting down-stream ventral visual processing.

**Figure 3.**
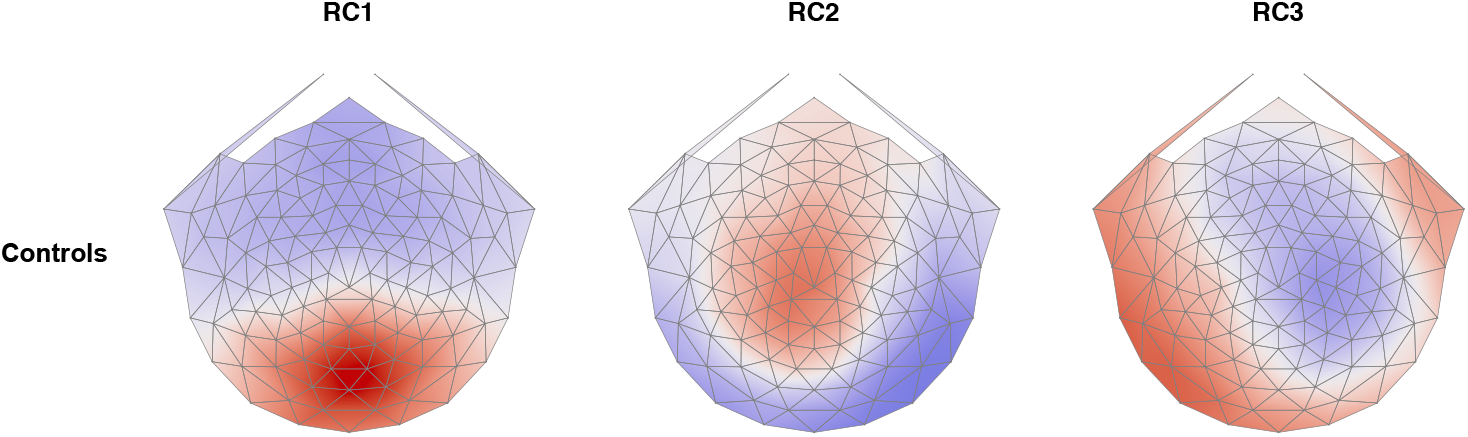
Scalp topography of the first three RCs for controls. The color-maps are on the same arbitrary scale. For each topography, north is towards the nasion, south is towards the inion.

### Ignoring dependency inflates performance

Individuals left and right eyes ‘MD and VEP responses are correlated (See Figure 1). If one wishes to use both eyes in a classification analysis, it is important to account for this dependency structure to restrain overly-optimistic classification performance (Rosset & Tibshirani, 2019). We have accomplished this by keeping eyes belonging to an individual in the same fold in k-fold cross-validation. The effect of respecting this dependency structure is demonstrated in Table 1 where we report the classification performance metrics. This table contains the rates for 12 classifiers in total (2 algorithms x 3 pathway combinations x 2 CV techniques). First, we draw attention to the rows falling under the “paired CV” and “ignore-pairing CV”. Here, the classifiers that ignored the pairing falsely “out-perform” the classifiers that considered pairing; the increase in AUC averaged across all 6 classifier columns is approximately 5%. Note, this better performance in the pairing-ignored case is because, in a given CV fold, the training dataset is not independent from the test dataset due to some participants having eyes (which are correlated) in both datasets at once. Classification sensitivity is particularly important in glaucoma, as early detection of the disease is key for effective treatment. The pairing-ignored sensitivity was inflated by approximately 3%, on average. Overall, paired CV classification performed moderately well, with AUCs of 0.7, on average, meaning around 70% of cases were correctly identified.

**Table 1:** Summary of classification results for both the logistic regression and random forest, for both pairing strategies, and for all pathway-biasing stimuli.

|  | Logistic regression |  |  | Random Forest |  |  |
| --- | --- | --- | --- | --- | --- | --- |
|  | Both | On | Off | Both | On | Off |
| Paired CV |  |  |  |  |  |  |
| Accuracy | 0.685 | 0.637 | 0.678 | 0.651 | 0.616 | 0.647 |
| Sensitivity | 0.768 | 0.689 | 0.797 | 0.786 | 0.809 | 0.795 |
| Specificity | 0.579 | 0.571 | 0.529 | 0.503 | 0.417 | 0.511 |
| AUC | 0.707 | 0.67 | 0.701 | 0.72 | 0.694 | 0.723 |
| Ignore pairing CV |  |  |  |  |  |  |
| Accuracy | 0.719 | 0.656 | 0.703 | 0.684 | 0.656 | 0.7 |
| Sensitivity | 0.802 | 0.751 | 0.808 | 0.834 | 0.816 | 0.817 |
| Specificity | 0.614 | 0.536 | 0.571 | 0.531 | 0.482 | 0.574 |
| AUC | 0.761 | 0.683 | 0.728 | 0.792 | 0.742 | 0.787 |

### OFF-features yield small performance improvement

While the recent isolated-check VEP literature has focused on using ON-pathway features for glaucoma classification, recent work has suggested that the OFF-pathway may be more vulnerable to glaucomatous insult. Indeed, under our conditions of measurement, classification using only OFF pathway features performed better than classifiers using ON pathway features. This is shown in Table 1, where we will focus on the pairing-CV classification. Here, for Logistic regression and Random Forest, AUCs were an average of 2.3% higher for OFF-pathway features. For Glaucoma, early detection is key for preventative therapy, so the sensitivity of classification is of particular importance. Interestingly, in the random forest, sensitivity was very slightly better for the ON-pathway (though specificity was poorer), but for the logistic regression sensitivity was considerably reduced in the ON-pathway, while the OFF-pathway performance was maintained. Overall, in consideration of the AUCs, we report a small improvement in performance for the OFF-pathway, to the extent that the OFF-pathway performance alone is on-par or better than the performance yielded when both ON and OFF features are considered.

The better performance of OFF-pathway features may be reflected in the features retained by the lasso or the feature importance in the random forest. As such, we evaluate the importance of features in two ways. First, for our logistic regression, we look at the features retained by the elastic-net feature selection. Second, we look to the Random Forest, where we inspect the mean decrease in Gini score for the top 15 features. In both cases, we look only at the outcome for the paired-eye CV when both ON- and OFF-features are considered for classification. The matrices in Figure 4 show the features that were retained by the elastic net (dark cells are retained features). The left-most plot is for ON-features, and the right-most for OFF-features. Within each plot, RCs are represented on the y-axis, while phase (Cos and Sin theta), amplitude, and harmonic are represented on the x-axis. First, the OFF pathway features retained (15) outnumbered the ON-pathway features retained (12) - a small difference of approximately 11% (relative to the total number of features retained). The ON-pathway features are scattered amongst the matrix, while the OFF-pathway features appear more concentrated into RC1, particularly into the phase-related components, consistent with the notion that the phase of the OFF pathway is effected by Glaucoma (Norcia et al., 2022).

**Figure 4.**
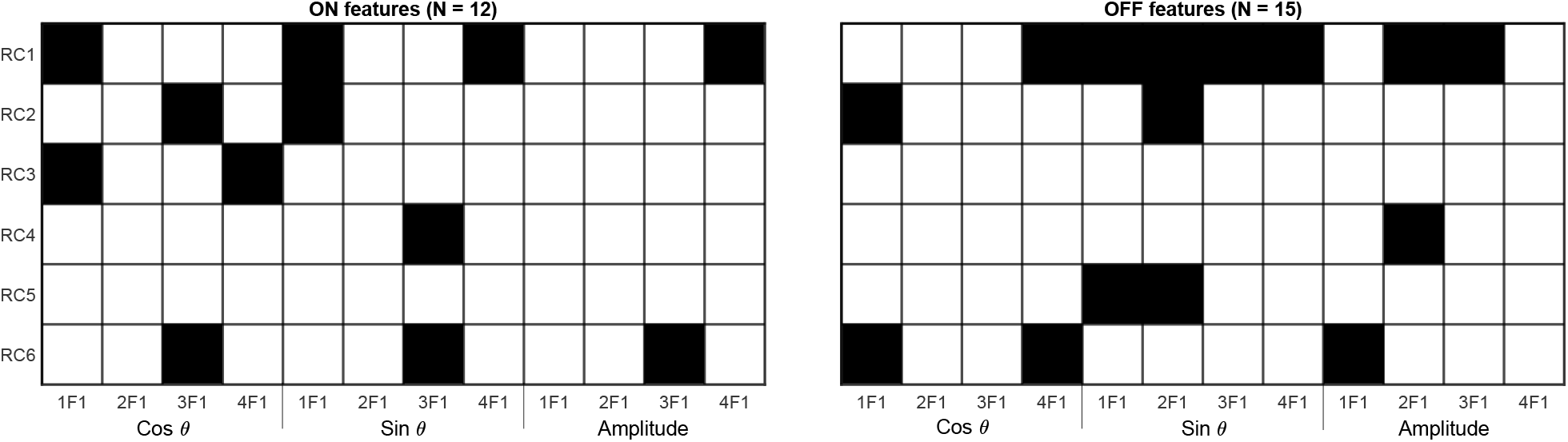
Plot of feature selection matrix after training using paired-eye CV. Separate plots are provided for ON (left panel) and OFF (right panel) pathway features. Dark cells indicate retained features. Fifteen OFF-related features are retained compared to 12 ON-related features.

The results of our logistic regression show that OFF features out-number ON features when using the elastic-net feature selection. However, this says nothing of the hierarchy of feature importance. To investigate the relative importance of features that are driving classification, we turn to the Random Forest. In Figure 5 we show the mean decrease in Gini score for the top-15 features. First, it is noteworthy that the most important feature is an OFF-feature, the OFF amplitude of the 2nd harmonic in RC1, followed by an ON-feature (the cosine phase of the fundamental for RC1). In consideration of all top fifteen features, eight are OFF features and six are ON-features, but age alone is a better predictor than two of the ON-features. Furthermore, OFF-features tend to cluster towards the top of the variable importance plot, such that 4 of the top 5 features are OFF features.

**Figure 5.**
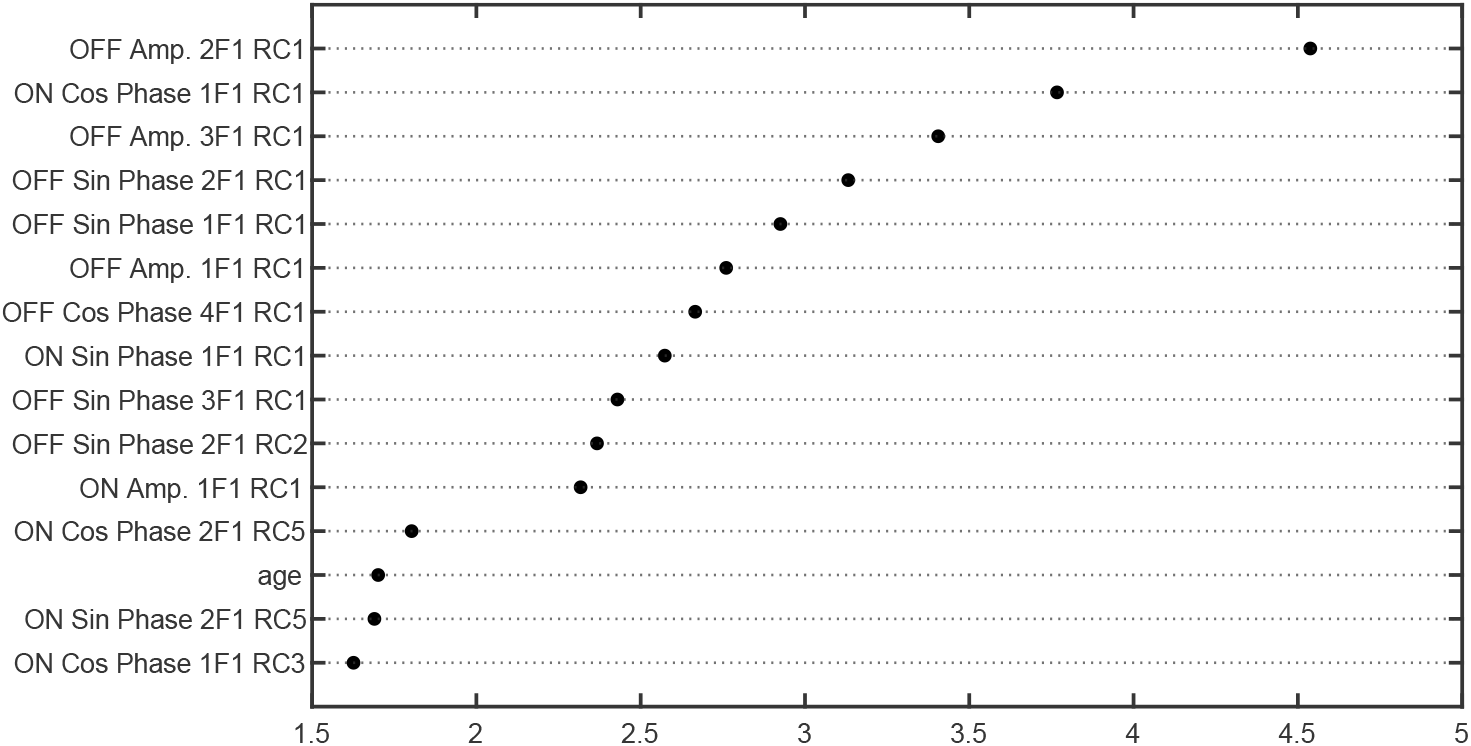
Gini feature importance plot for random forest classifier for the top 15 features

## Discussion

Our results on individual patient classification for glaucoma indicate that SSVEP features derived from OFF-pathway-biased saw-tooth stimulation are better able to distinguish patients from controls than ON-pathway-biased features. This is manifests in higher accuracy and AUC values for classification on the basis of OFF-pathway biasing features over ON-pathway biasing features, the inclusion of more OFF features in the classifier and the higher ranking of OFF features in feature importance for classification. Beyond reinforcing differential effects of glaucoma on the ON vs OFF pathways, the present results suggest the potential utility of the SSVEP approach to individual patient prediction in the clinic.

### Relative utility of OFF-vs ON-response features

The central goal of our study is a comparison of the relative utility of ON-vs OFF-pathway derived response features in discriminating patients with glaucoma from controls. As noted, we find a small improvement in accuracy for OFF-derived features over ON-derived features (a difference of 3% AUC) and little benefit of combining features (0.5% AUC). OFF-derived features are more frequently retained in the classifier (15 vs 12 features, respectively), and our analysis of feature importance from the Random Forest classifier indicated that OFF-features are generally of higher value for classification. This is in-line with recent reports that indicate that the OFF-pathway is preferentially damaged by glaucoma in humans (Norcia et al., 2022) and in mouse (Daniel et al., 2018; Puyang et al., 2017), and suggests that further exploration of this OFF-pathway vulnerability would be fruitful for biomarker development. However, the clinical utility if the SSVEP *per se* is still undetermined. Our best-performing classifier was only moderately effective, producing AUCs of approximately 70%. For comparison, recent classification analysis of OCT data (Shin et al., 2021) and Humphrey visual-field images (Akter et al., 2022) has yielded AUCs and F1 scores in excess of 90%. How might classification rates be improved for the SSVEP? We collected relatively little data from the two eyes over the contrast polarity manipulation. This is because this protocol was embedded in a larger set of stimulus conditions designed to optimize and examine other study variables. This limited the available signal to noise ratio which may limit the absolute classification rates. The extent to which SSVEP-based classification is limited by SNR should be examined, as this can be improved in future work by simply collecting more trials, or using higher contrast stimuli (although, the effect of glaucoma on high-contrast visual perception is not well understood).

### Comparison with previous studies of VEP Glaucoma classification

Despite our novel inclusion of both eyes in training and having many more EEG features than previous work, our overall classification rates are lower than those reported previously from the icVEP literature. In the paired-eyes analysis, our best AUC score was 72% (for the OFF-features random forest classifier) and the best accuracy obtained was 68% (for both ON and OFF features in the logistic regression. For comparison, Zemon et al. (2008) using 10 Hz sinusoidal stimulation at 10% contrast, found classification accuracy was higher for decrements than increments (91% vs 73%, respectively), but at 15% contrast, accuracy was higher for increments than decrements (94% vs 82%). That is, their worst accuracy was higher than our best accuracy. Following the work of Zemon and colleagues, Fan et al. (2018), in a much smaller sample (37 patients, 26 controls), reported a maximum accuracy of 77%, while X.-W. Chen and Zhao (2017) obtained accuracy’s as high as 90% - both of these reports only used the ON-biasing icVEP. In terms of AUCs,L. J. Xu et al. (2017), reported AUCs as high as 80%, for ON-pathway biasing icVEPs, and Kolomeyer et al. (2020) achieved even better rates, reporting AUCs of 85% - 90%. What might explain the reduced performance we have obtained? Aside from the aforementioned limitation of a low number of trials in the present work, one possibility is that previous reports provide overly optimistic metrics of performance. Here, we have used 10-fold cross-validation to produce robust performance estimates and minimise over-fitting. The performant binary classifiers from the aforementioned work have chosen a univariate SSVEP SNR-based classification threshold based on the receiver-operator-characteristic curve derived from their enitre dataset. Then, they find a threshold that produces the best performance in the same dataset. This is equivalent to training and testing a classifier on the same data, which increases the probability of model over-fitting. This improves classification rates for a specific dataset, but the performance may not generalise to new data. The authors may consider reanalysing their data using some form of train-test splitting (as we have done with cross-validation). Regardless of our reduced absolute classification rates, which are subject to variables such as the amount of data collected, we find that accuracy is higher for OFF than for ON-pathway features derived from saw-tooth stimulation. Given the numerous other differences between our approach and that used in the isolated check literature, future studies would do well to continue to perform direct comparisons of results from different forms of ON-vs OFF-pathway selective stimulation. Finally, assessment of sensitivity to pre-perimetric damage and obtaining relative measures of test-retest reliability, treatment effects, and progression would also be of interest to determine the ultimate utility of the approach.

## Conclusions

The present results, in combination with previous work in human glaucoma and animal models of glaucoma, suggest that the OFF-pathway is more vunerable to glaucoma, and thus carries more diagnostic information than the ON-pathway for this pathology. The extent to which the SSVEP is useful for in-clinic diagnoses is still undetermined, but we show that further investigation of OFF-pathway specific biomarkers is merited.

## Data Availability

All data produced in the present study are available upon reasonable request to the authors

## Acknowledgments

The authors gratefully thank support from the Glaucoma Research Foundation, the National Eye Institute (P30-EY026877 and R01-EY030361-01) and Research to Prevent Blindness, Inc. Vladimir Vildavski developed the instrumentation used in this study.

